# Phase angle clusters in bioimpedance: An alternative to body mass index

**DOI:** 10.1101/2024.01.02.24300705

**Authors:** Ma Jesús Fuentes Sebio

## Abstract

**Background:** The “Body Mass Index” (BMI) or “Quetelet Index” is the most widely used tool to diagnose the degree of obesity. Anyone can calculate it, with no other tools than a scale and a measuring rod. However, its accuracy in predicting body fat percentage is low. The aim of this study is to find an alternative to BMI that is more reliable, accessible and easily applicable in daily clinical practice.

**Methods:** Using the K-means method (an unsupervised classification algorithm), we performed a *cluster* analysis of the two phase angles obtained with bioimpedance analysis (BIA) of 641 women with different health status. BMI, age, diseases, treatments and any other data other than the phase angle values of the participants were not taken into account.

**Results:** The *clusters* generated by the K-means algorithm do not coincide with the BMI categories, nor with the predetermined division of individuals into healthy and pathological.

The K-means clustering algorithm identified new patterns that provide information on the greater or lesser predisposition of different individuals to suffer from diseases, taking as a reference their pathological peers in the same *cluster*.

**Conclusions:** The categories generated by the K-means algorithm based on the phase angles obtained by BIA classify individuals according to their health status independently of other variables such as age or BMI.

## 1 Introduction

The Belgian astronomer and mathematician Adolphe Quetelet (1796-1874) was ahead of his time in the creation of databases for study and evaluation. One of his best known analyses was the relationship between the weight and height of individuals (what we now call the Body Mass Index or BMI) contained in his book “Sur l’homme” [66]. Based on population statistics, he formulated the aforementioned index also called Quetelet’s index in 1836. His interest in statistics led him to search for mathematical laws that could be adapted to social phenomena, delving into subjects such as education, birth-mortality and even delinquency. What is surprising is that at no time did he suggest that BMI could be applied as a diagnostic value [66]. It was Ancel Keys in 1972 who proposed it to classify people with obesity [14]. It was not the only marker designed to define weight status, although it was the most widely used.

The simplicity of the “Body Mass Index” formula led to its widespread use [6], constituting a very intuitive marker [7] of the level of body fat deposits. Its formula is as follows:

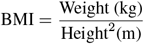

What is striking is that BMI does not reflect the size of the adipose tissue [3] and is incapable of distinguishing changes in the muscle mass, bone and water compartments [5, 15]. This means that we should not be surprised by the report of metabolic disturbances in women with normal BMI with a medium-high percentage of fat [37]. This incongruence was also found in cardiovascular pathologies, in which individuals with low BMI presented greater complications than their counterparts with a higher BMI [38]. Patients with the same diagnosis of heart failure, hypertension or peripheral vasculopathies [8, 10, 9] with low or even normal BMI had worse prognosis than those with higher BMI. Cardiovascular risk, based on the Framingham risk score, is associated with an increase in BMI. What is surprising is that this only predicts 30 % of the cases of the aforementioned risk [4]. This shows that the categories delimited by BMI are not adequate when classifying individuals and even less so when applying them to clinical diagnosis and prognosis.

The optimal BMI range in some medical circles was established between 20-25 kg*/*m^2^, considering that in this range of values, the risk of mortality is the lowest. However, the risk of diabetes increases from a BMI of 20 kg*/*m^2^, so relying on such an inaccurate index is a big mistake [40].

BMI does not reflect fat concentration or location. Specifically, **visceral fat**, unlike subcutaneous fat, is found surrounding the internal thoracic and abdominal organs, including the heart [58, 59]. Excess visceral fat is a predisposing factor for inflammatory pathologies, insulin resistance, and dyslipidemia [62, 63]. In 1970, Florey indicated that the BMI, as well as other similar indexes, should not be used to classify individuals according to their degree of adiposity. However, and despite the criticism, the BMI has been widely used in recent years [42, 41].

There are several technologies for calculating body composition: computed tomography (CT), magnetic resonance imaging (MRI), dual X-ray absorptiometry (DXA), hydrometry, ultrasound, isotopic dilution with deuterium or tritium and many others. All of them are limited by their cost, complexity or side effects [47, 48, 49, 51]. On the other hand, bioimpedanciometry (BMI) is the technique that is gaining more and more followers day by day due to its ease of use, safety, low cost and relatively good reliability, although without reaching the effectiveness of other more sophisticated and precise techniques [28, 20].

**Bioimpedance** is an inexpensive and non-invasive technique. It measures the difficulty of the alternating current passing through the different body structures. It is a very useful and practical tool for both clinical and research purposes [20]. It consists of two components: resistance (R) and reactance (Xc) [29]. The resistance is the opposition of body tissues to the flow of electric current. The reactance reflects the dielectric properties of cell membranes, real biological capacitors that generate a delay in the time of electric current circulation [50].

**Phase angle** (PhA) is the most robust parameter of bioimpedance, reflecting cellular integrity and hydration status [52, 53]. It is defined as the geometric angular transformation of the ratio of reactance to resistance and can be calculated with the following formula [21, 22]:

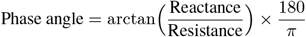

Any variation in the reactance and resistance values is reflected in the values of the phase angle which represents the duration of the interval from when the electricity passes through the membranes until the voltage changes [43, 44].

Phase angle is a marker of cellular wellness [16, 27]. It is a good reflection of muscle mass and physical activity [18, 19]. High PhA values indicate a good state of health while low values are related to pathologies [32]. Several studies reported a direct relationship between the phase angle and pathologies such as cancer [23], AIDS [22] or other chronic diseases [16, 17]. This makes PhA a powerful tool for monitoring the clinical progression of a given pathology or a change in health status [25].

**Clustering** encompasses a group of unsupervised techniques whose purpose is to create groups or patterns from the data provided. The observations within each group are similar to each other and different from those in the other groups. Unlike supervised machine learning techniques, clustering does not require a training sample because, being an “unsupervised” method, the process ignores the response variable that indicates to which group each observation belongs (if such a variable exists).

Clustering is used in the most diverse and varied fields of science with a variety of applications [36, 2, 26]. The **k-means algorithm** is one of the best known and most widely used clustering techniques [54]. It is fast and easily applicable to large data sets and it is used in machine learning techniques and pattern recognition [60, 61]. It is based on the random search of a set of points called **centroids** (average value of the points of that *cluster*), grouping the different objects in their nearest centroid [55] based on a distance metric which is usually the “Squared Euclidean distance”.

## 2 Objective

The aim of this study is to find a reliable alternative to BMI to classify individuals based on phase angle to predict increased vulnerability to metabolic disease. The aim is to group individuals into new categories with clinical significance.

## 3 Methodology

Observational, retrospective, cross-sectional study including bioimpedanciometric records of 641 white female participants aged 19 to 74 years with a BMI between 17.90 and 56.30 kg*/*m^2^.

The multi-frequency BIA equipment TANITA MC-980MA with 8 electrodes [56] was the bioimpedance meter at 50 kHz frequency. The measurements were carried out in an upright position with an ambient temperature of 21 °C according to the following requirements:

- No eating or drinking in the last 4 hours.
- No exercise in the last 24 hours.
- Not taking diuretics.
- No alcohol intake in the 8 hours prior to the measurement.

From the data collection, two groups were created with female individuals. One group consists of individuals with no treatments or diseases. The second group comprised women with pathologies such as hyperlipidemia and/or insulin resistance treated (or not) with hypocholesterolemia and/or oral antidiabetics. Individuals with neoplasms, cardiac, hepatic or renal insufficiency as well as persons under treatment with antihypertensives and corticosteroids were excluded ^1^. People medicated with levothyroxine, anxiolytics, contraceptives and proton pump inhibitors were not excluded from the study. Table 1 details the number of people with or without treatment as well as the type of treatment.

**Table 1:** Participant groups and treatments.

| Treatments | Number of individuals |
| --- | --- |
| None | 588 |
| Oral antidiabetics | 5 |
| Hypolipidemic agents | 23 |
| Oral antidiabetics and hypolipidemic agents | 3 |
| Contraceptives and hypocholesterolemic | 1 |
| Proton pump inhibitor and hypocholesterolemic | 5 |
| Anxiolytics and hypocholesterolemic | 10 |
| Levothyroxine and hypocholesterolemic | 6 |

The **K-means** method, an unsupervised and non-deterministic classification algorithm [45] that groups the data into *k* groups based on the minimum sum of distances between each object and the centroid of its group or *cluster*, is applied to the data set. It is a very agile and versatile algorithm that optimizes the similarities of the data within each *cluster* [46].

Applying the K-means method, four non-overlapping *clusters* of phase angle values were generated, taking into account the minimum sum of distances between the object and the centroid of the *cluster*, as shown in Figures 1 and 2. The number of *clusters* was defined based on the “elbow method”, which calculates the average distance from the centroid to all *cluster* points. That is, it analyzes the percentage of variance explained as a function of the number of *clusters* [64].

**Figure 1.**
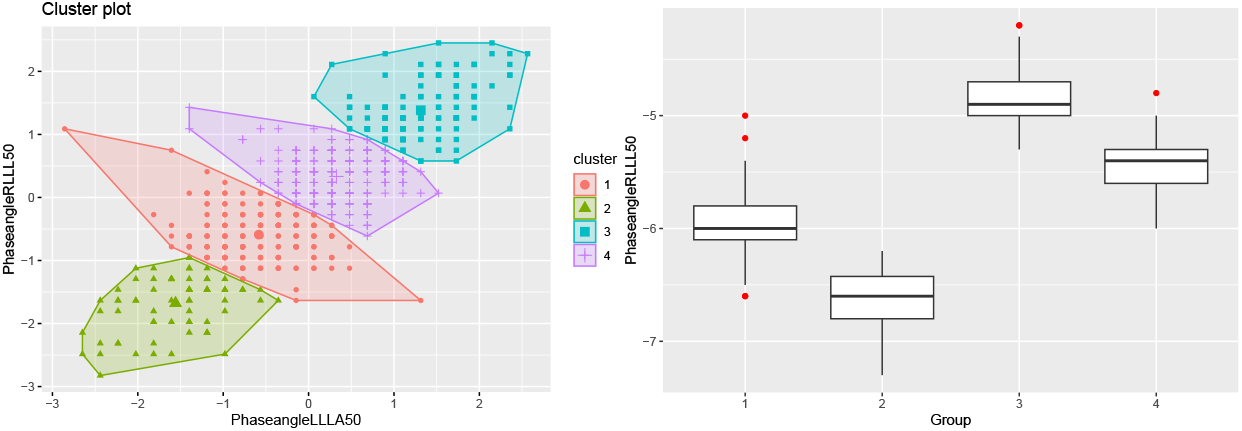
Plots of the four resulting clusters.

**Figure 2.**
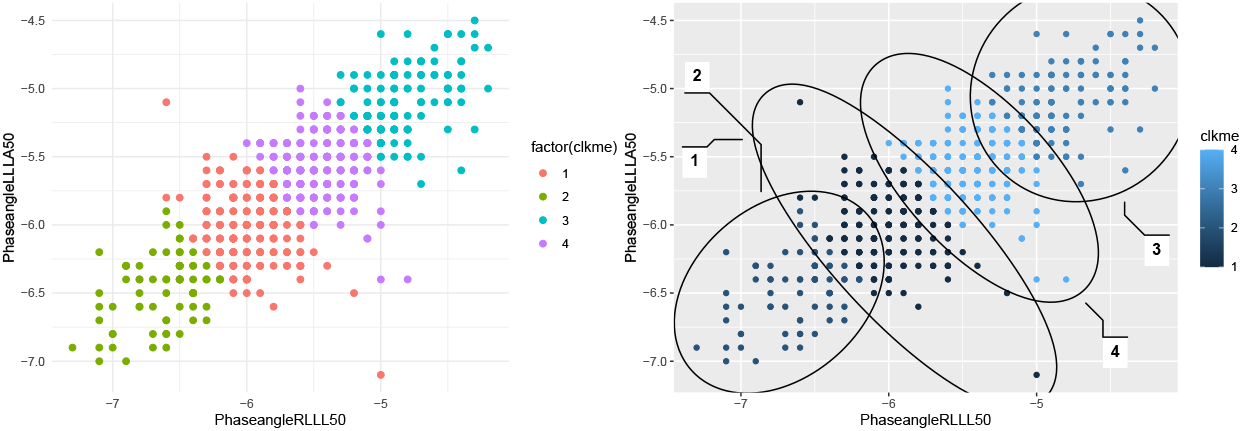
: Different representations of the four *clusters*.

## 4 Cluster Analysis

If we analyze the *clusters* obtained from lowest to highest, we see that number 2 is the one with the lowest phase angles followed by groups 1, 4 and 3. To assess the differences between them, we studied the health conditions as well as the treatments of the pathological individuals in each *cluster*:

### Cluster 1

It consists of 209 individuals, 195 of them without health problems and 14 with pathologies, including, in addition to hyperlipidemia and type II diabetes mellitus, multi-organ pathologies and autoimmune diseases that curiously are not found in the other *clusters*. It could be said that this is the **most pathological group** from the metabolic point of view. We see it detailed in Table 2.

**Table 2:** Cluster 1.

| N° | Medication | Pathologies |
| --- | --- | --- |
| 1 | Simvastatin and anxiolytics drugs | Hyperlipidemia |
| 2 | Simvastatin, Insulin and oral hypoglycemic agents | Pluripathological |
| 3 | Metformin | Polycystic ovarian disease |
| 4 | Atorvastatin | Hyperlipidemia and intestinal pathology |
| 5 | Simvastatin and anxiolytics drugs | Hyperlipidemia and untreated rheumatoid arthritis |
| 6 | Hypolipidemic agents and Levothyroxine | Hyperlipidemia and hypothyroidism |
| 7 | Metformin | Type II Diabetes and autoimmune pathology |
| 8 | Hypolipidemic agents | Hyperlipidemia and liver disease |
| 9 | Simvastatin and anxiolytics drugs | Hyperlipidemia and total hysterectomy |
| 10 | Simvastatin | Hyperlipidemia, renal pathology and polycystic ovarian |
| 11 | Atorvastatin and Levothyroxine | Hyperlipidemia and hypothyroidism |
| 12 | Simvastatin and Levothyroxine | Hyperlipidemia and hypothyroidism |
| 13 | Simvastatin and oral Contraceptive | Hyperlipidemia and polycystic ovarian |
| 14 | Atorvastatin | Hyperlipidemia |

### Cluster 2

This group is constituted by 74 individuals of which 3 are pathological. It is the *cluster* with the **lowest phase angle** and, curiously, the 3 pathological patients suffer from **muscular pathology**. One of them with chronic elevation of the enzyme Creatinphosphokinase (CPK) and the other 2 suffer from osteomuscular pathology under study. The summary of these 3 pathologic cases can be seen in Table 3.

**Table 3:** Cluster 2.

| N° | Medication | Pathologies |
| --- | --- | --- |
| 1 | Metformin | Elevated CPK muscle enzyme y polycystic ovarian |
| 2 | Simvastatin | Hyperlipidemia and muscular pathology |
| 3 | Rosuvastatin | Hyperlipidemia and muscular pathology |

### Cluster 3

It consists of 105 healthy individuals and 17 pathological individuals. This is the group with the **highest phase angle values**. There were no significant pathologies except hyperlipidemia and 3 individuals with type II diabetes, as shown in Table 4.

**Table 4:** Cluster 3.

| Nº | Medication | Pathologies |
| --- | --- | --- |
| 1 | Simvastatin | Hyperlipidemia |
| 2 | Rosuvastatin | Hyperlipidemia e hysterectomy |
| 3 | Insulin and oral antidiabetics | Type II Diabetes |
| 4 | Oral antidiabetics | Type II Diabetes |
| 5 | Simvastatin and Ezetimide | Hyperlipidemia |
| 6 | Simvastatin | Hyperlipidemia |
| 7 | Simvastatin | Hyperlipidemia |
| 8 | Simvastatin | Hyperlipidemia |
| 9 | Simvastatin and anxiolytics drugs | Hyperlipidemia |
| 10 | Metformin | Type II Diabetes and hysterectomy |
| 11 | Simvastatin and Gemfibrocil | Hyperlipidemia |
| 12 | Simvastatin and Anxiolytics drugs | Hyperlipidemia |
| 13 | Pitavastatin | Hyperlipidemia and polycystic ovarian |
| 14 | Simvastatin and Levothyroxine | Hyperlipidemia |
| 15 | Simvastatin and anxiolytics drugs | Hyperlipidemia |
| 16 | Rosuvastatin and anxiolytics drugs | Hyperlipidemia |
| 17 | Simvastatin | Hyperlipidemia |

### Cluster 4

This *cluster* is formed by 217 women without pathologies and without treatments, 19 with hyperlipidemia and 2 individuals of this group who suffer from type II diabetes as shown in Table 5.

**Table 5:**
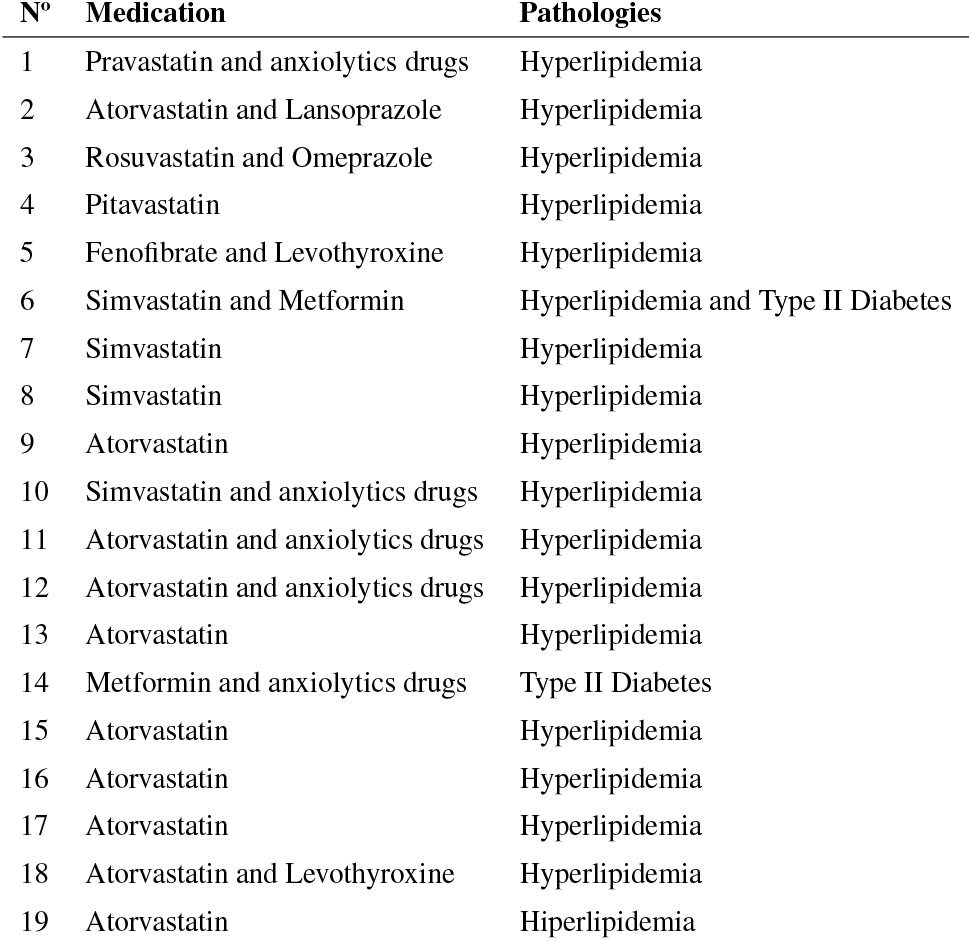
Cluster 4.

| Nº | Medication | Pathologies |
| --- | --- | --- |
| 1 | Pravastatin and anxiolytics drugs | Hyperlipidemia |
| 2 | Atorvastatin and Lansoprazole | Hyperlipidemia |
| 3 | Rosuvastatin and Omeprazole | Hyperlipidemia |
| 4 | Pitavastatin | Hyperlipidemia |
| 5 | Fenofibrate and Levothyroxine | Hyperlipidemia |
| 6 | Simvastatin and Metformin | Hyperlipidemia and Type II Diabetes |
| 7 | Simvastatin | Hyperlipidemia |
| 8 | Simvastatin | Hyperlipidemia |
| 9 | Atorvastatin | Hyperlipidemia |
| 10 | Simvastatin and anxiolytics drugs | Hyperlipidemia |
| 11 | Atorvastatin and anxiolytics drugs | Hyperlipidemia |
| 12 | Atorvastatin and anxiolytics drugs | Hyperlipidemia |
| 13 | Atorvastatin | Hyperlipidemia |
| 14 | Metformin and anxiolytics drugs | Type II Diabetes |
| 15 | Atorvastatin | Hyperlipidemia |
| 16 | Atorvastatin | Hyperlipidemia |
| 17 | Atorvastatin | Hyperlipidemia |
| 18 | Atorvastatin and Levothyroxine | Hyperlipidemia |
| 19 | Atorvastatin | Hiperlipidemia |

It is noteworthy that the last two *clusters* (3 and 4) do not present major differences between them in terms of treatment and pathologies. However, *cluster* 3 is the one with the highest phase angle values. To help reveal this difference, the **visceral fat** indices provided by the application of the bioimpedancemeter were compared, taking into account that a good correlation was reported between visceral fat measured by Computed Tomography (CT) and Bioimpedancemetry [57].

As we can see in Table 6, *cluster* 4 is the one with the highest values of visceral fat, which means a greater predisposition to suffer from metabolic pathologies. This means that *cluster* 3 would be the one with the best metabolic health conditions.

**Table 6:** *Cluster* 3 and 4 visceral fat index analysis.

| *Cluster* | *Mean* | *Standard deviation* |
| --- | --- | --- |
| 3 | 7.098 | 3.269 |
| 4 | 7.275 | 3.139 |

According to the above, the k-means *cluster* classification divided the database into 4 groups according to their phase angle. The resulting *clusters* with the most pathological individuals are found in the lower part of the graph where the phase angles are lower. This means that the healthy partners in these *clusters* should be monitored for their potential predisposition to muscle or metabolic diseases. Conversely, individuals with a higher phase angle would be in a more optimal state of health.

If we analyze the BMI (based on the classification of Table 6) of each *cluster*, we can see that in all of them, there are individuals with different BMI values, as we can see in Figure 3. This means that the classification carried out by k-means does not coincide with that of BMI.

**Figure 3.**
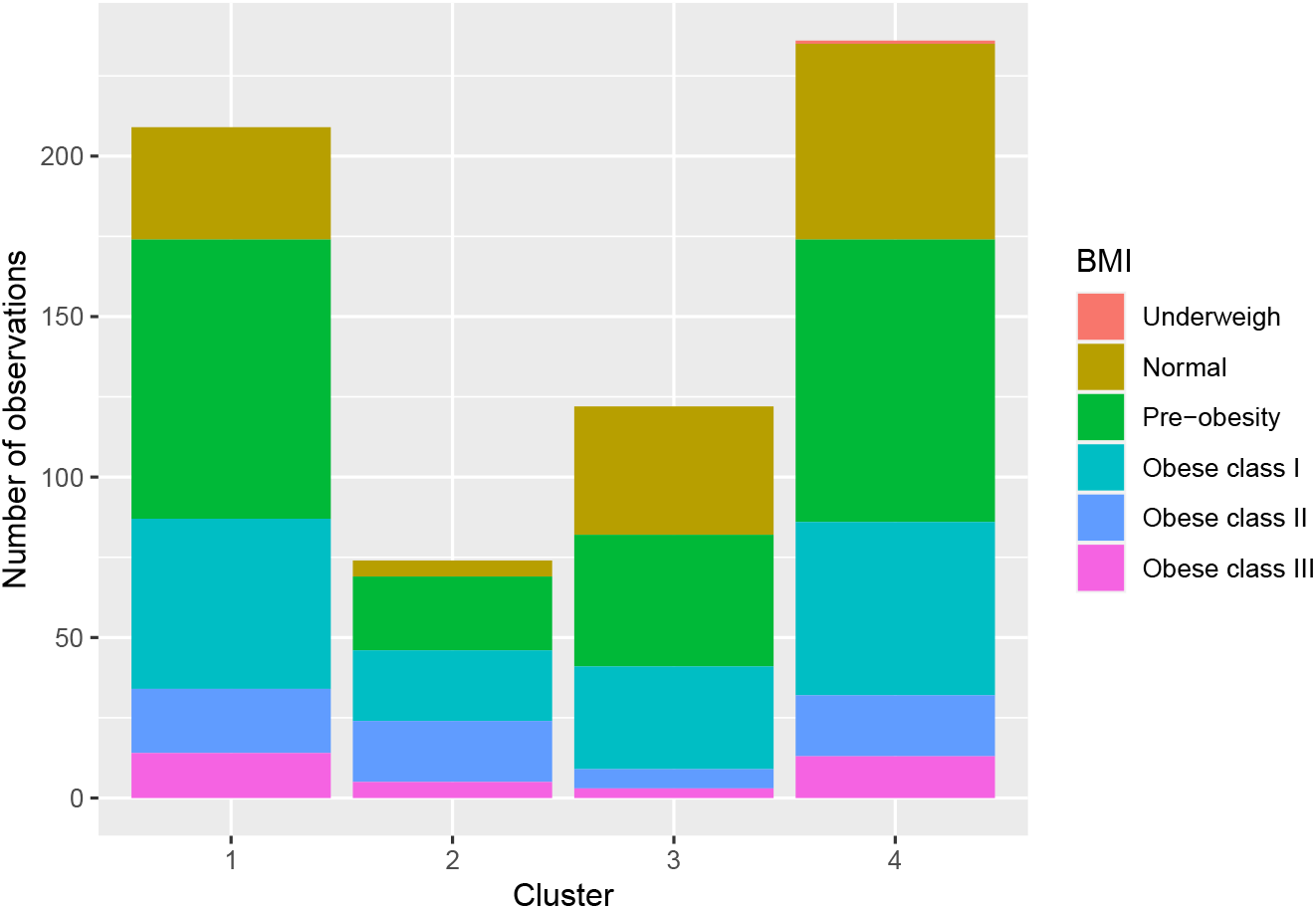
: *Cluster* chart according to the BMI classification reflected in Table 7.

## 5 Discussion

The Body Mass Index (BMI) has a long history of use due to its speed and accessibility in diagnosing the degree of obesity. However, it does not achieve the degree of fidelity necessary to discriminate individuals with risk factors associated with an increase in their fat component. The BMI does not reflect the distribution or concentration of adipose tissue, a situation that leads to overlooking individuals with high visceral fat, a highly determinant factor in the development of metabolic pathologies. Thus, individuals with the same BMI but with different fat distribution present disparate clinical manifestations because the increase in energy reserves does not have the same health consequences in subcutaneous adipose tissue as in visceral adipose tissue [33, 34, 35].

**Table 7:** BMI classification [65].

| **BMI** | **Weight status** |
| --- | --- |
| <18.5 | Underweigh |
| 18.5 - 24.9 | Normal |
| 25.0 - 29.9 | Preobesity |
| $\geq 30.0$ | Obese |
| 30.0 - 34.9 | Obese class I |
| 35.0 - 39.9 | Obese class II |
| $\geq 40$ | Obese class III |

Novel techniques are needed to differentiate those individuals with risk factors associated with their increased body mass. BMI does not distinguish between alterations in body composition such as increased muscle mass in athletes or muscle loss in the elderly.

Phase angle is one of the most robust parameters provided by bioimpedance. It reflects the state of hydration and cell mass [27] as well as being a marker of cell well-being [16].

In this research study, a **k-means clustering** of a database of the values of two phase angles obtained by BIA of people with and without metabolic pathologies was performed using the k-means algorithm. No classificatory variables that could distort the investigation were introduced to the algorithm.

The k-means generated four clearly differentiated *clusters* based on the values of their phase angles. Those with the lowest phase angles included individuals with muscle pathology, which raises suspicions about the integrity of the musculoskeletal system of the supposedly healthy individuals in the same category.

The individuals in the next *cluster* do not have such low phase angles, but it could be considered the most complicated, metabolically speaking, since the most pathological individuals are concentrated in this *cluster*.

The last *clusters* (three and four) present similar characteristics although their phase angles are different, so the visceral fat index determined by BIA was used to evaluate the latter, which was found to be higher in the group of individuals with lower phase angles, a situation that predisposes them to the development of metabolic pathologies.

The categories performed by k-means reflect the health status of the individuals, which makes phase angle a powerful predictor of metabolic diseases as well as muscle pathologies.

It would be advisable to extend the study to individuals of other sexes and with different pathologies in order to glimpse clues that will help the clinician when approaching the therapeutic alternatives available. This study has limitations such as the lack of standardization of the different bioimpedance devices available on the market as well as the need to carry out similar studies with larger and more varied samples.

It would be interesting to create several lines of research in different fields of medicine to establish a reference pattern of phase angle values to be applied in daily practice and thus catalog groups of individuals by their predisposition to suffer certain pathologies. Cut-off points that will help us to classify in a first consultation the need or not to delve into diagnostic evaluations.

## 6 Conclusion

The categories generated by the K-means algorithm, based on phase angle values obtained by BIA, classify individuals according to their health status, constituting a much more reliable, non-invasive and relatively simple alternative to BMI for daily clinical practice.

## Data Availability

All data produced in the present study are available upon reasonable request to the authors.

## Supplementary material

## Footnotes

1 They produce changes in total body water value.

